## Supplemental Materials for "The Tumor-Associated Calcium Signal Transducer 2 (TACSTD2) oncogene is upregulated in pre-cystic epithelial cells revealing a new target for polycystic kidney disease"

#### Supplemental Material Table of Contents

**Supplemental Table 1**: Differentially expressed genes at P6 (Excel Spreadsheet).

**Supplemental Table 2**: Differentially expressed genes at P10 (Excel Spreadsheet).

**Supplemental Table 3**: Overlap of Differentially expressed genes at P10 and ADPKD (Excel Spreadsheet).

**Supplemental Figure 1**: Small RNAseq of Pkd2 experimental kidneys vs control at P10 identifies microRNAs with known association in PKD.

**Supplemental Figure 2**: Differentially expressed mRNAs in pre-cystic and early cystic mouse kidneys.

**Supplemental Figure 3**: *Tacstd2* expression in cystic epithelium.

**Supplemental Figure 4**: Tacstd2 expression in cystic mouse renal epithelium.

**Supplemental References**

Supplemental Figure 1

**A**

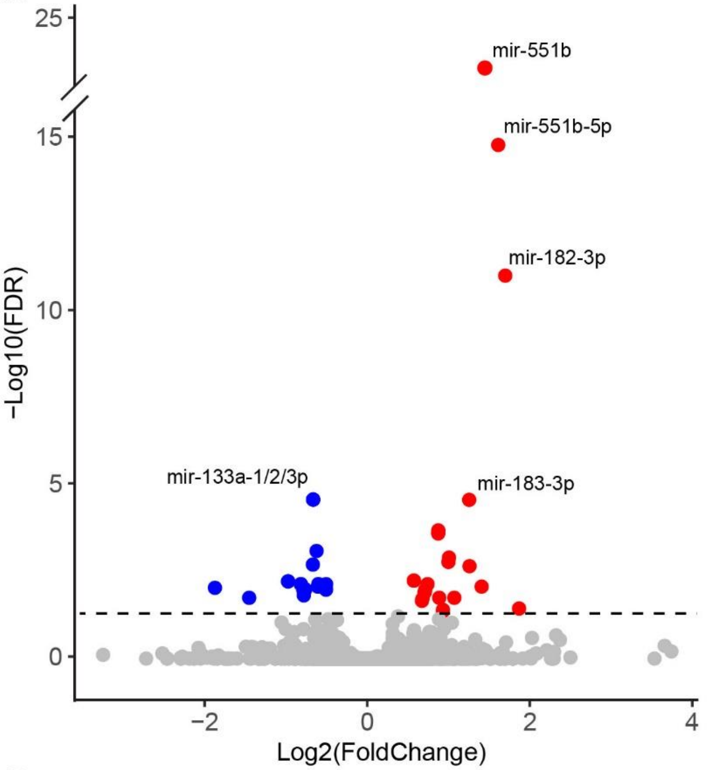


**B**

| miRBase ID | log2 Fold Change | FDR | Gene Family or Cluster | PKD Literature |
|---|---|---|---|---|
| mmu-miR-18a-3p | 0.889 | 0.018 | mir-17-92 cluster | Patel et al (2013) |
| mmu-miR-92b-3p | 0.710 | 0.012 | mir-17-92 cluster | Patel et al (2013) |
| mmu-mir-92b | 0.709 | 0.012 | mir-17-92 cluster | Patel et al (2013) |
| mmu-miR-182-3p | 1.702 | <0.001 | mir-182 cluster | Pandey et al (2011); Woo et al (2017) |
| mmu-miR-183-3p | 1.258 | <0.001 | mir-182 cluster | Pandey et al (2011); Woo et al (2017) |
| mmu-mir-182 | 1.010 | 0.001 | mir-182 cluster | Pandey et al (2011); Woo et al (2017) |
| mmu-miR-182-5p | 1.002 | 0.002 | mir-182 cluster | Pandey et al (2011); Woo et al (2017) |
| mmu-mir-183 | 0.745 | 0.007 | mir-182 cluster | Pandey et al (2011); Woo et al (2017) |
| mmu-miR-183-5p | 0.717 | 0.010 | mir-182 cluster | Pandey et al (2011); Woo et al (2017) |
| mmu-mir-96 | 0.683 | 0.019 | mir-182 cluster | Pandey et al (2011); Woo et al (2017) |
| mmu-miR-96-5p | 0.677 | 0.022 | mir-182 cluster | Pandey et al (2011); Woo et al (2017) |
| mmu-mir-21a | 0.935 | 0.039 | mir-21 family | Lakhia et al (2016) |
| mmu-miR-21a-5p | 0.934 | 0.043 | mir-21 family | Lakhia et al (2016) |
| mmu-miR-222-3p | 0.577 | 0.006 | mir-221 cluster | Ben-Dov et al (2014) |
| mmu-mir-222 | 0.577 | 0.006 | mir-221 cluster | Ben-Dov et al (2014) |
| mmu-miR-551b-5p | 1.615 | <0.001 | mir-551 family | Ben-Dov et al (2014) |
| mmu-mir-551b | 1.445 | <0.001 | mir-551 family | Ben-Dov et al (2014) |
| mmu-miR-133a-3p | -0.662 | <0.001 | mir-133 family | Ben-Dov et al (2014) |
| mmu-mir-133a-1 | -0.662 | <0.001 | mir-133 family | Ben-Dov et al (2014) |
| mmu-mir-133a-2 | -0.662 | <0.001 | mir-133 family | Ben-Dov et al (2014) |
| mmu-mir-338 | -0.620 | 0.001 | mir-338 family | Ben-Dov et al (2014) |
| mmu-miR-338-5p | -0.665 | 0.002 | mir-338 family | Ben-Dov et al (2014) |
| mmu-mir-488 | -0.813 | 0.007 | mir-488 family | Pandey et al (2011) |
| mmu-miR-488-3p | -0.813 | 0.007 | mir-488 family | Pandey et al (2011) |

**Figure S1. Small RNAseq of *Pkd2* experimental kidneys vs control at P10 identifies microRNAs with known association in PKD.**

(A) Volcano plot showing P10 differentially expressed microRNAs (Omiras). Colored dots indicate FDR <0.05, red are upregulated and blue are downregulated in Experimental vs. Control. Plot was cropped for better visualization of data.

(B) Table indicates microRNAs described in (A) that were previously cited in literature with relation to polycystic kidney disease. Citation refers to the individual microRNA or its related family or cluster. Red are upregulated and blue are downregulated in our results.

### Supplemental Figure 2

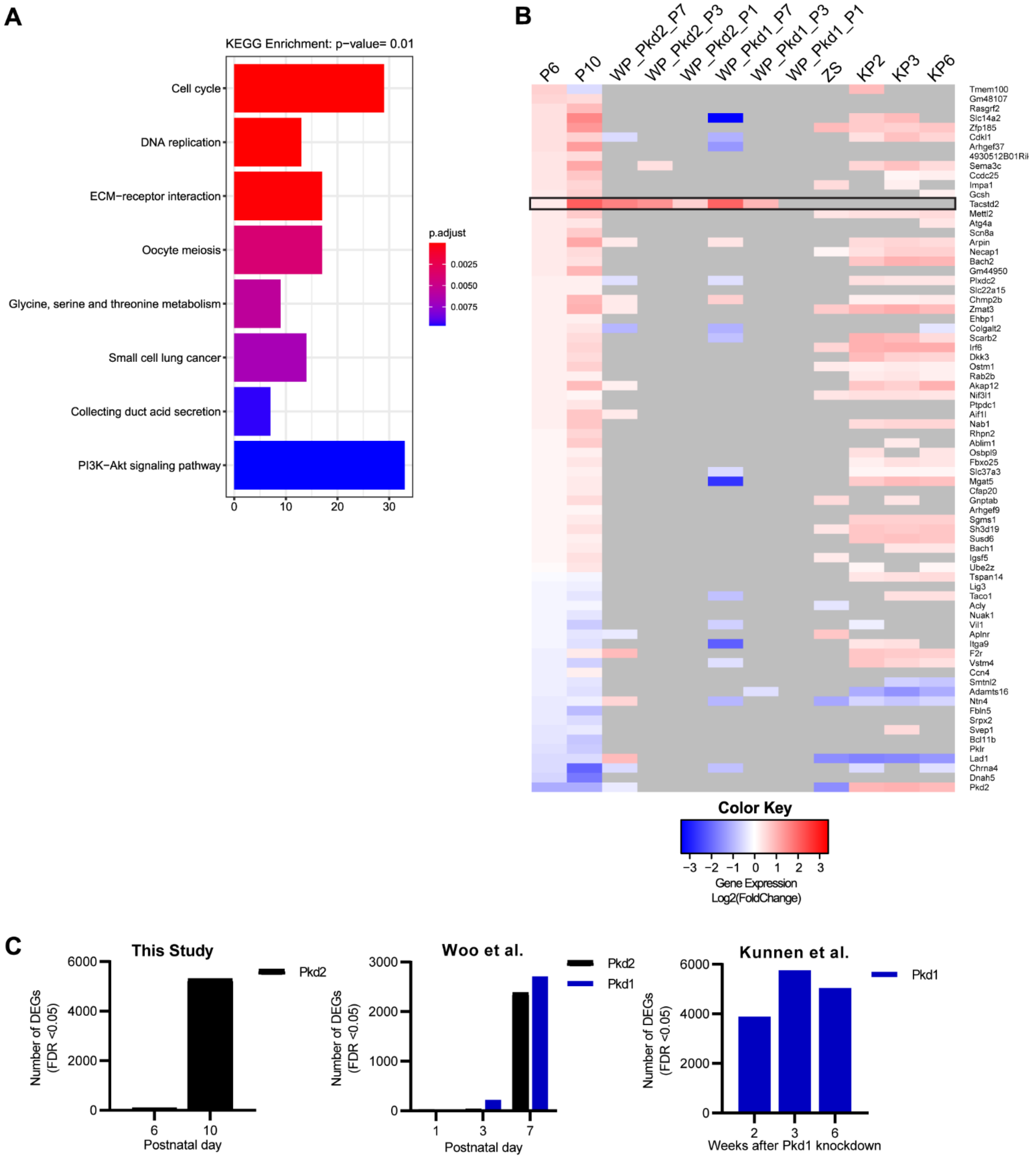

**Figure S2. Differentially expressed mRNAs in pre-cystic and early cystic mouse kidneys.**

(A) KEGG pathways found to be enriched for P10 differentially expressed mRNAs (DEGs).

(B) Heatmap comparing differential gene expression for cyst initiation candidates (CICs) in our data (P6, P10) and published pre-cystic mouse data [WP_Pkd2_P7/3/1, WP_Pkd1_P7/3/1 [1], ZS [2], KP2/3/6 [3]. *Tacstd2* identified with label and black box.

(C) Barplots showing the number of differentially expressed genes (False discovery rate < 0.05) identified in data from the described studies. Our study: P6 and P10 following Pkd2 deletion. Woo et al.: P1, P3, and P7 following Pkd1 or Pkd2 deletion. Kunnen et al.: Weeks 2, 3, and 6 following Pkd1 deletion. Pkd1 models represented by blue bars). Pkd2 models represented by black bars.

### Supplemental Figure 3

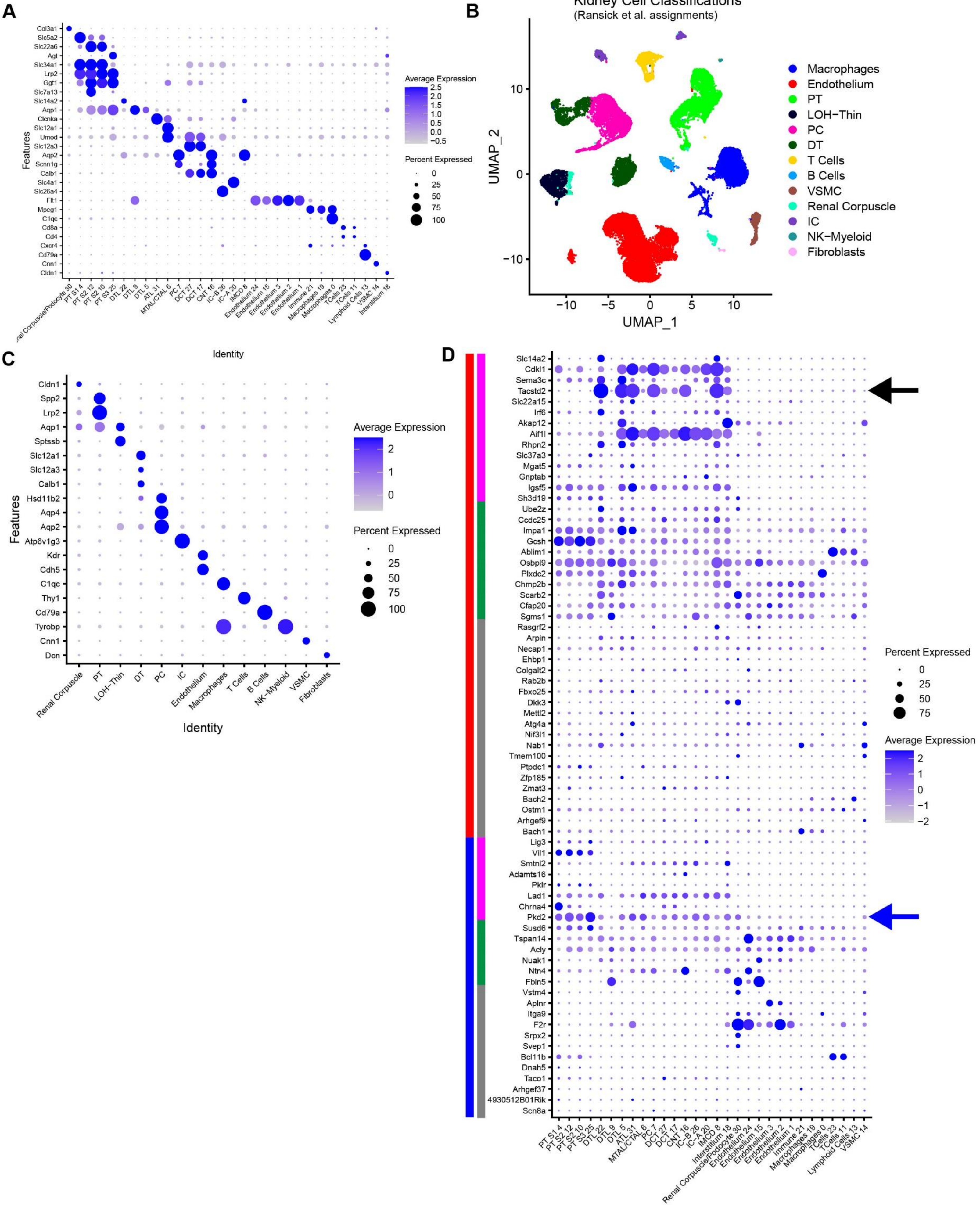

**Figure S3. *Tacstd2* expression in cystic epithelium.**

(A) Dotplot of marker genes supporting cell type assignments in Fig 3E. Markers according to Chen et al. [4]. Dot radius represents percent of cells in cluster expressing the gene. Opacity scale indicates average gene expression level within expressing cells. Row normalized.

(B) UMAP from Fig 3E with cell type assignments as labeled by Ransick et al. [5].

(C) Dotplot of marker genes used to annotate Fig S3B. Dot radius represents percent of cells in cluster expressing the gene. Opacity scale indicates average gene expression level within expressing cells. Row normalized.

(D) DotPlot visualizing the localization of CICs to the murine kidney cell atlas. Dot radius represents percent of cells in cluster expressing the gene. Opacity scale indicates average gene expression level within expressing cells. Row normalized.  Main bars (far left) represent increased expression (red) or decreased expression (blue) in our data; sub bars (left) represent epithelial restricted expression (magenta), broad expression (green), and stromal/indeterminate (gray). Black arrow points to *Tacstd2*, an epithelial cyst initiating candidate. Blue arrow points to PKD2.

Supplemental Figure 4

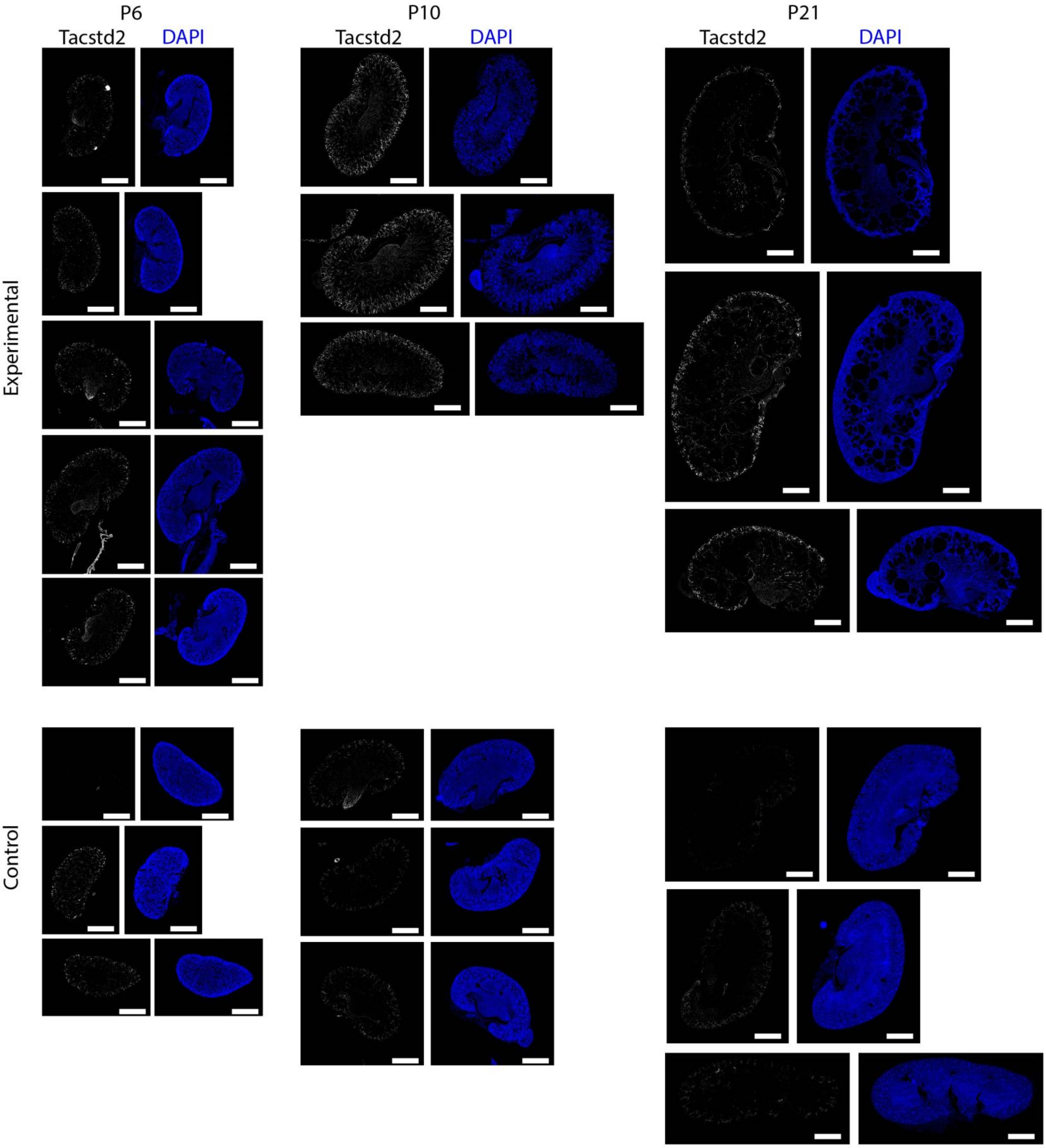

**Figure S4. Tacstd2 expression in cystic mouse renal epithelium.**

(A) All scans used for quantification described in Fig 4B. Scale bars 1mm.
